# Mitigating the identity and health threat of COVID-19: Perspectives of Middle-Class South Asians living in the UK

**DOI:** 10.1101/2021.03.30.21254537

**Authors:** Kristin Hanson, Emma O’Dwyer, Sharmistha Chauduri, Luiz Gustavo Silva Souza, Tushna Vandrevala

## Abstract

The recognition and representation of BAME community as “high risk” of Covid-19 in the UK presents both a health and an identity threat to this ethnic group. This study employed thematic analysis to explore response to these threats as related by a sample of thirteen middle class members of the South Asian community. This work advances both health and identity psychological theory by recognising the affinity between expressions of health efficacy and identity. Our findings identify South Asian intragroup stigmatisation and commonalities that have implications for the promotion of health behaviour and health communications for minority groups.

---

As the COVID-19 pandemic unfolded in the spring of 2020, the United Kingdom (UK) public became aware of the unequal way in which the country’s population was being affected by the disease. Initially reported as an “equalizer” in terms of morbidity and mortality, the SARS-CoV-2 (COVID-19) pandemic is now considered to discriminate, inflicting a disproportionate burden of illness and death across BAME communities in the UK (Aldridge et al., 2020; Office for National Statistics, 2020a). Analysis of deaths involving COVID-19 by ethnicity for England and Wales published by ONS in 2020 showed that taking account of age and other socio-demographic characteristics and measures of self-reported health and disability at the 2011 Census, the risk of a COVID-19-related death for people of Black ethnicity was 1.9 times higher than for those of White ethnicity. Mortality amongst a Bangladeshi and Pakistani ethnic group was 1.8 times more likely than in the White ethnic comparison group (Office for National Statistics, 2020a). More recent statistics from London suggests that London’s Asian populations have been worst affected during the second wave of the pandemic, followed by Black communities (Fenton, 2021). Media reports also consistently highlighted the disproportionate scale of the impacts of COVID-19 on ethnic communities, often treating the risk for ethnic communities as being homogeneous. Although tracking the effect of the virus by ethnicity is important for the purposes of identifying inequalities and interventions, the grouping together of the BAME community as “high risk” introduces a context in which these groups are presented with increased threat based on their BAME ethnicity. This paper seeks to explore responses to this threat from the perspective of more advantaged members of the BAME, particularly identifying how this threat has manifested in participants’ assertions of health efficacy and identity.

In the UK, the largest non-White ethnic group is comprised of people of South Asian (7.0%, Bangladeshi, Indian, and Pakistani) ethnicities (Office for National Statistics, 2013). Within this South Asian ethnic group—like other broad ethnic groups—there are native country cultural differences, differences in religion, and differences in the historical association with the adopted country. In the UK, there are also wide disparities in the socio-economic status within this group. In 2019, Pakistani and Bangladeshi people earned 16% and 15% less than White people, respectively. In contrast, those from Indian ethnic groups earned 16% more than White ethnic groups (Office for National Statistics, 2020b). These differences are enough to contest the homogenous grouping of British South Asians let alone the broad grouping of BAME in the UK pre-pandemic. British South Asians may occupy what (Jaspal and Cinnirella, 2012: 507) refer to as ‘threatening positions’ even in times in which actual identity threat is not present due to difficulties in reconciling their ethnic and national identities. Indeed, the negotiation of ethnicity and nationality for this group is the subject of a number of studies (Jaspal and Cinnirella, 2013; Robinson, 2009; Vadher and Barrett, 2009). However, social change, such as that precipitated by the COVID-19 pandemic and the related risk to the BAME community, may reactivate the need to negotiate these roles (Cinnirella, 1997). The context of the current study therefore provides a unique opportunity to gain insight into how these negotiations interface with health threat responses.

## COVID-19 Threat in the BAME Community

In the context of the COVID-19 pandemic and the related media coverage of risk to the BAME community, an individual in this group was likely to have been facing at least two threats: being involuntarily categorised by ethnicity in a negatively evaluated group, and being at a higher risk of disease. BAME groups have a higher COVID-19 incidence and mortality due to greater presence of co-morbidities (such as insulin resistance, Type 2 diabetes mellitus, cardiovascular disease, central obesity and essential hypertension, and vitamin D deficiency) with these conditions being linked to poor outcomes for COVID-19 (Onder et al., 2020). Socio-economic challenges, such as being poorer, material deprivation, high-risk occupation/ front-line public-facing jobs, the location of residence, household composition and overcrowding in housing, with extended family in multi-generational households, with implications for transmission from younger to older and more vulnerable household members (Dhillon et al., 2020; Mamluk and Jones, 2020).

Not only is it clear that the virus is a greater threat to their personal health, but this group is also being marked out as different from the rest of the UK population, potentially presenting a threat to their ethnic and national social identities. When an individual is categorised in a social group against their will or there is a threat to the group’s value, a social identity threat can occur (Branscombe et al., 1999; Breakwell, 1986). Branscombe et al. (1999: 40) have indicated that involuntary categorisation “may be particularly threatening in a context where that group membership implies poor ability or performance” such as may be the case if the higher risk in the South Asian community is construed as a consequence of non-compliance with public health guidance.

## Identity and Efficacy

Both health psychology and social psychology suggest that individuals will employ a variety of strategies when confronted with health and identity threat. In many of the health psychology theories used to predict behavioural change (REFS), and for a given threat, individuals consider various physical and psychological resources and options (Lazarus and Folkman, 1984) such as efficacy beliefs (D’Amico et al., 2013). Self-efficacy is an individual’s belief in their own ability to exercise influence over their life and accomplish desired tasks (Bandura, 1977). In the context of health, self-efficacy refers to the individual’s belief in their ability to enact health behaviours, as well as their belief about the effectiveness of that behaviour (Evangeli et al., 2016; Garcia, 2016; Holden, 1992). Self-efficacy is not a personality trait, but is dependent on both psychological and social factors that comprise a set of self-beliefs (Bandura, 2006). Despite its strong predictive ability, self-efficacy is problematic, particularly when considering minority populations. For example, recent evidence on ethnic communities have concluded that South Asian and other ethnic communities are less likely to espouse self-efficacy in the context of health threat (Hendy et al., 2019; Vandrevala et al., under review). In addition, previous research has shown that the expression of self-efficacy is entwined with the external environment for migrants (Hendy et al, 2019, Vandrevala et al, under review). For example, people from ethnic communities were less likely to be efficacious if they felt that the health behaviour was less likely to be valued or benefit the health of their families or communities. Migrants from the South Asian continent were less likely to displace self (individual) self-efficacy as they came from collective societies, and felt misunderstood or not welcome by the health system (Hendy et al., 2019).

Recent studies have highlighted the importance of group and institutional efficacy resources in response to threat, particularly in the context of societal risk, such as COVID-19 (Cho et al., 2020; Cho and Kuang, 2015). Of direct relevance here is Cho et al.’s 2020 paper that researched the stigmatisation of the Asian community in the US. In this work, the researchers theorised that self-, group- and institutional efficacy would guard against stigmatisation of an outgroup during COVID-19. Group efficacy is the belief that groups work together to achieve intended outcomes and institutional efficacy concerns the confidence in the effectiveness and efficiency of organizations, which draws on the belief that societal institutions are considered fair, trustworthy, and predictable (see Cho et al. 2020). Cho and colleagues found that self-efficacy was not related to stigmatisation, group efficacy was associated with decreased stigmatisation, and increased institutional efficacy (in their study, the government) was related to increased stigmatisation. In the face of high threat, perceptions of low collective efficacy and/or institutional efficacy may lead to maladaptive coping responses, which may include stigmatisation of others.

The social psychological literature on identity elaborates this link between self-efficacy and identity. Specifically, feelings of efficacy can be seen as integral to Breakwell’s (1986) identity process theory (IPT). IPT recognises self-efficacy (defined as feeling in control of one’s life) as an objective of a person’s belief about their identity. This identity principle, along with self-esteem (feelings of personal worth), distinctiveness (from others or other groups), and continuity (across time and situations) are seen as the primary motives of identity construction (Breakwell, 1986, 1993, 2001). In this way, this literature also recognises that self-efficacy and identity construction are reciprocally linked. Specifically, Breakwell notes that “the individual may engage in the exercise of self-efficacy” (1986: 102); and that the process of identity construction can provide the individual with feelings of control and competence (Jaspal and Breakwell, 2014). This ‘exercise of self-efficacy’ constructs self-efficacy as an agentic response to threat, a resource that can be created by the individual, one that is an *enacted* part of identity construction. Viewing the relationship between self-efficacy and identity construction in this way highlights the unique opportunity to evaluate efficacy and identity in the current context.

Response to categorisation threat is dependent on a number of elements, particularly the status of the group (Doosje et al., 1995, 1999) and the level of an individual’s commitment to the group (Branscombe et al., 1999; Ellemers et al., 2002). Those whose identification with the threatened BAME group is low may invoke a number of responses. They may emphasise the heterogeneity of the South Asian group and stress their own personal qualities. They may also seek individual mobility by disidentifying with the negative social category and emphasizing their membership in a higher status group, thereby ensuring a positive self-image. In the context of the current study—one in which health threat is coupled with identity threat—we would expect these expressions of identity to be intertwined with expressions of efficacy.

## The Current Study

The current study sought to explore how response to threat is managed within a community that has been identified as being at particular risk from COVID-19. In particular, we focus on members of the community for whom this segregation is likely to be an identity threat due to the transition in social status that it represents: South Asians who are established, middle-class immigrants. In interrogating this response to what is both a health and identity threat, we seek to understand to what extent efficacy is employed and how these maps on to identity construction and health mobilisation.

## Method

This study examined representations of the COVID-19 threat through semi-structured online synchronous interviews. A qualitative approach was selected to best capture the complexity of this context.

### Participants

We recruited a sample of 13 participants for this study using opportunistic and snowballing sampling methods (Ritchie et al., 2003), limiting the number of participants to allow an in-depth exploration of their representations related to the South Asian community and COVID-19 (Ritchie et al., 2003). Employing a convenience sample as a basis for understanding perspectives in a population has the limitation of generalisability to a wider group; in addition, a sample in which a high level of motivation is required to complete the study task (the interview), may over- or under-represent certain perspectives. All participants have been resident in the UK for more than ten years and are considered to have or have had white-collar employment. The majority of the participants were of Indian origin and of Hindi faith. None of the participants had contracted COVID-19 before the date of the interview. Demographics of the group are indicated in Table 1 below.

Potential participants were identified through the third author’s personal network; they were solicited initially telephone and if they indicated interest, were asked to return their informed consent via e-mail. Upon receipt of consent, a mutually agreeable interview time was arranged. No participants were considered to have specialist medical or health knowledge. The research received a favourable ethical opinion from Kingston University, London.

### Procedure and Materials

The data were collected in May 2020, approximately one month after the first national ‘lockdown’ was initiated in the UK. The semi-structured interview schedule included approximately 12 open-ended questions that focused on participants’ representations of COVID-19 risk, barriers, and facilitators in the South Asian community. Typical questions included “What do you think are the health concerns for people in your community during this pandemic and why?”, “How do you think your, the South Asian community, has specifically been affected by the coronavirus?”. By allowing the participants to discuss COVID-19 in terms of the community in which they have been placed by the government and media, the interview schedule aimed to grant participants the freedom to manage this identity. The third author, who shared ethnic and migrant identities with the participant group, conducted the interviews. Interviews were conducted in English using Zoom and were most commonly completed after 60-75 minutes.

### Data Analysis

The interviews were audio recorded and transcribed by the interviewer. Interview transcripts were then imported into MAXQDA 2020, a qualitative analysis software application, for organisation and coding. A form of thematic analysis was chosen to explore the data due to its epistemological and analytical flexibility (Braun and Clarke, 2006). An inductive approach was taken, ensuring a bottom-up analysis of the data rather than one driven by particular theoretical objectives. We were, however, alert to the means by which the participants understood and managed the risk asserted.

All analyses were conducted by the first author. Prior to initial coding, the data corpus was read and re-read. Initial thematic codes were then generated using a line-by-line approach, ensuring that all of the data were given equal attention. With a view to capturing both the representations of health risks, facilitators, and barriers as well as how the participants positioned themselves in relation to the threat, coding identified both semantic and latent items. In this initial coding, codes were assigned to the entire collection of data, participant by participant. The codes were reviewed and discussed within the author group, identifying the key themes related to threat management. Codes were then pruned to identify and consolidate themes, and these themes were reviewed based on their relevance to the research question. The themes were then named, defined, described, and interpreted. Primary themes are discussed in an integrated fashion below.

## Findings

Through thematic analysis, three themes were developed: (1) Contesting the definition of the South Asian community, (2) Enacting self-efficacy: Taking responsibility to mitigate personal health risk, and (3) Constructing the integrated immigrant identity. These themes collectively create a narrative for the management of the identity and health threat.

### Contesting the Definition of the South Asian Community

We found that our participants managed their group efficacy by resisting categorisation in the South Asian community, aligning themselves with the British majority, and recasting themselves as members of an alternative group. Few of the participants accepted their position in speaking for the ‘South Asian community’ without question; they instead often reconfigured the definition of their community to meet their understanding of their identities. On the whole, participants declared little difference between the South Asian and wider communities based on ethnicity alone.

Resisting the category of South Asian was manifest explicitly by some participants in denial of community knowledge: “*I don’t have any direct, direct communication with the, with the people, you know, Asians or South Asians*” (P3), or “*I don’t have any contact with Bangladeshi community here, so I don’t know, I don’t know…I know nothing about them*” (P11).

An alternative approach of distancing taken by the participants was to assert the lack of difference between the South Asian and the majority white population in the UK: “*So these days, you know, South Asian, Western or British or English or whoever is all the same*” (P9). Some were opposed to health messaging by the government that classified people into groups “*It says, you know, the government, so as they say, there are black family, white family! They do not have to say that!*” (P6). The resistance to the South Asian category offers the participants the opportunity to redefine their ‘community’ in accordance with their perceptions. Important social groups for these participants included their neighbourhood, religious groups, and family.

In a positioning that is similar to the ‘we are all the same’ distancing from the South Asian label identified above, some participants considered their community to be one comprised of their neighbourhood, regardless of ethnicity: “*You see, in my road, I have a mixed population*” (P12). This community group provided significant support for some during lockdown. Participant P9 credited her neighbourhood group efficacy: *“Luckily, we have good neighbours, they are helping. But if nobody were there to help, then you had it!”*.

But for many, the primary source of social support was their family unit, a group efficacy that was evident in participants’ recounting of how older generations benefited from being close to younger members of their family, facilitating compliance: “*I think the elderly generation, probably got the messages from their children, most of them live with the family, you know*.” (P6). Younger generations were relied upon to deliver food and provided social interaction. This multi-generational living arrangement that is a feature of South Asian culture was an acknowledged tension:

> *And the chances are of more affected, because they live in extended families. They live together. The parents live together, in some cultures, the children look after the parents. The children have to travel, so when the children come home, they’re mixing with the parents who are elderly. So that affects the parents as well. Because that is their culture. But when you take Western culture, they are on their own. And also, they’re on their own, they are also struggling nobody to help. So whether you are Western or you are Asian, it is the same problem, but here they are really always worried that they shouldn’t be going near their bed. And at this pandemic, that is an advantage, for the family living together*. (P9)

The participant referred to the sharing of households by multiple generations as both dangerous and protective. In her perception, it was dangerous if family members did not adopt protective behaviours to avoid the infection of older members of their household. It was protective if they adopted such behaviours and the younger family members provided care and solidarity. This description of the risk and benefit of the multi-generational living culture positions the tradition in contrast to the risks of the stereotypical Western culture living in nuclear families.

By acknowledging the negative impact that the closure of religious and community facilities has had on the group, the importance of religious groups was communicated by a number of participants, both in a lament for closed centres: *“I think to some extent there has been dis-integration of community, partly because of closure of the most of the community”* (P5), and in the role of community leaders, particularly religious leaders as key catalysts in the dissemination of health messaging and the promotion of compliance during the pandemic: *“I would say different clubs or who run or religious places can inform their people that what has been happening and how they can take the precautions”* (P1). The centrality of religion was, like multi-generational living, seen as an integral part of South Asian culture: *“The way that culture in India is, people, right from childhood, they have been born and they’ve been developed with some contextual spirituality in some form…I think in many contexts is imbibed in people who have come from there”* (P2).

The participants asserted their version of the groups that were important to their identities: their neighbours, their family, and their religious groups. In light of this identity re-definition, we next examine how participants managed the COVID-19 health and identity threat in this context.

### Enacting Self-efficacy: Taking Responsibility to Mitigate Personal Health Risk

Participants identified that the South Asian community may be at greater risk due to underlying disease such as cardiovascular disease and diabetes, “*Well the concern is, that many of the South Asian community people have got underlying healthcare conditions-it is diabetes, it is high blood pressure, it is asthma and, uh, old age related other problems*.” (P13).

The perceived susceptibility of the South Asian community to the disease was attributed to elements that were under their perceived behavioural control or self-responsibility:

> *So, you got to be self-disciplined, not because government is telling you; government may say whatever it is, but it is your own responsibility, for your own health, safety and security*. (P13).

Some attributed the increased risk to physical disadvantages related to not being in their native country, including hypothesised decreased immune-system effectiveness due to the lack of exposure to disease “*poor country people have lot more immunity*” (P12) and Vitamin D deficiency due to the combination of the lack of sun and greater skin melanin:

> *we come from these tropical countries and all of this are lack of sunlight over here. And a Vitamin D deficiency also causes certain of the things like depression like lower immunity and then probably those are one of the reasons*. (P4)

The majority of participants attributed the cause of underlying risk of COVID-19 to the relative lack of focus on particular health habits, such as diet and exercise in the South Asian community. In these discussions, participants drew a distinction between themselves and other segments of the South Asian population; new immigrants and older generations were cast as groups with this lack of health discipline:

> *I think the most important thing is the diet. Yeah. I saw most of them suffer from diabetes, high blood pressure. Lack of exercise. I used to go I see lots of Asian ‘kakas’ (uncles), elderly, you know. What they do is, I mean, they go to the gym*… *They just sit in the jacuzzi* …*being lazy, in the gym*. (P6)
>
> *Well, I think our, our food, I am not the person, but our generation’s food is much more healthier. Food is much more healthier than people who recently arrived from India, for example*. (P5)

In the first passage, the participant refers to the South Asian community as an outgroup (them) and contrasts themselves as one who uses the gym properly in opposition to the older generation. P5 refers to ‘our food’ but is creating a group that is specific in generation and UK residency. In these expressions we begin to note how identity is constructed to manage health risk. The contrast between recent and more established immigrants within the South Asian community was also implicit in participants’ health and hygiene knowledge: *“So the public is not much aware of the cleanliness in general*.” (P7), and “*I think the possibly the British people I feel has learned the art of, you know, keeping, keeping good health from childhood. It was always part of the curriculum and everything*.*”* (P2). In these contrasts, participants aligned themselves more with the British majority than recent immigrants from South Asia.

Throughout the above discussion, we have touched on the theme of heterogeneity in the South Asian community asserted by our participant group that underlies their response to the threat presented. In the next theme, we explore more fully this reaction to threat in terms of the participants’ collective and institutional efficacy that create their integrated immigrant identity.

### Constructing the Integrated Immigrant Identity

Our participant group managed threat by aligning their identity with the British majority group and distancing themselves from the heterogeneous South Asian group. The participant group’s assertions of heterogeneity included positioning certain immigrants (the poor and/or unintegrated) as more at risk for the virus and of the Muslim community as a source of COVID-19 restriction violations. The result is the construction of an integrated immigrant identity.

Invoking the lack of difference discussed in the first theme between the South Asian and the majority White group in the UK, participants generally felt that any non-compliance in the South Asian community mirrored non-compliance in the wider UK community and was due to wilful ignorance. COVID-19 messaging to the South Asian community was regarded as having been widely received, with efforts by the government cited, wide access to media, older generations being supported by younger, and community groups playing a role in getting the message to everyone.

> *I have not come to any community or people who do not understand. I think people are generally well informed. There is not a single individual who do not know by now, what is coronavirus and how harmful it is to the mankind. So the message is already there. It is the question is of habit of implementing*. (P13)

In this passage, we see a version of the self-responsibility identified in the previous theme. Coupled with this is the participant’s representation of the success of the government’s messaging. This endorsement of the government was apparent throughout the data in the participant group’s high level of trust in the UK government: “*There is a lot of trust and people believe in the government, you would get the little bit of dissent anyway you know, but there is trust*.” (5). There was acknowledgement of the unique nature of the problems being faced and the clear messaging was appreciated: “*What more can we ask?”* (P5).

This trust in UK institutions extended to the National Health Service (NHS – the publicly funded system of healthcare in the UK) as well. The overwhelming majority of the participants unreservedly felt that the South Asian community has equal access to NHS services. The following position from P4 was typical:

> *I think it should be okay for everyone. That’s a good part of NHS, you know, that’s how I feel fundamentally NHS is kind of equal and that’s what coronavirus is. Coronavirus is equal for everyone, that’s how it should be*. (P4)

Any racism that may exist for other ethnic minority groups was not perceived by this group to affect their ability to access healthcare. Although not asked about racism, participants were aware of [media/activist] charges of racism within the service and specifically discounted these: “*I heard that, but I have no first-hand knowledge or information, but I have not come across any of these in South Asian community saying about this*.” (P13). This high level of institutional trust may be derived from participants’ comparison to institutions in their native countries, from an alignment with UK values in the effort to conform to the majority, or from the South Asian community link with the NHS (“*Most of them are Asians*” [P9]) and with the government: *“Our finance minister, Rishi Shunak, Shunak is Shaunak, you know, meaning the teacher of the rishis (sages). But anyway, he is a Punjabi. He is quite sharp and bright*.” (P10). This alignment extended to British values and norms. The commonality was often underpinned by a civic British identity, as in the following passage wherein Participant 4 refers to the ‘law of the land’, and a trust in British institutions:

> *It’s as simple as this, whoever you are, South Asian, European or African or any Americans or anybody are the localities who live here, English. It’s only as simple as like we follow the law of the land. We are safe. Our community is safe and the government is also functioning happily for us*. (P4)

By invoking the British value of “law of the land” and a trust in the British government, P4 not only contests categorisation as a member of the South Asian community but asserts their identity as a British citizen. In particular, our group of participants employed the UK value of the ‘rule of law’ to contrast with non-compliant out-groups within the South Asian community, particularly Muslims. At the time of data collection, Muslims were celebrating Ramadan. Non-compliance in the Muslim community was attributed to the tendency of the group to prioritise religious law over the “law of the land” (P10), casting them as poor examples of British citizens. Non-compliance was also attributed to a perceived perspective of Islam to put their fate in the hands of Allah: *“The Ramadan is going on, I think they don’t care. …They think whatever god does will happen. I also think so, time to time, looking at them-that they let that be with Allah!”* (P12). This positioning puts the Muslim South Asians in contrast to the reasoned, self-responsible in-group that our participants defined in describing their self-efficacy above. Ignorance of how to align with UK norms was generally derided. This derision was not necessarily directed at recent or poor immigrants, but instead those who were considered to have not made substantial effort to integrate:

> *…they are here, you know, it was 30, 40 years back in this country. They have still not learnt properly, you know, the right sort of culture, the context in terms of language and you know, of, uh, those sorts of aspects of, you know, uh, uh, ways of making them aware of what’s going*. (P2)

Participants were keen to distance themselves from the unintegrated migrant who had failed to assimilate into British society “*And I usually say, don’t be an Indian in this you know*” (P5). These groups of unintegrated South Asians were drawn in contrast to the participants’ own group. In this distinction, participants made reference to how their own group had adapted and assimilated to integrate with British values and norms. P10 here references that even traditional women in their community had adopted wearing practical western clothing (trousers), as opposed to traditional sari and kurtas, as it was more suitable for the British weather.

> *now we are not all relying on God. We now know we have to help God to help us I went to a Guajarati community, a charity as such. I saw the lady there with trouser and shoe-Guajarati ladies. Because of the winter and cold things, they see it as hygienic. And they say, no, we have to do, we have to change it*. (P10)

Overall, the participants were keen to enforce their community efficacy, expressing that they felt their own community (however defined) was adhering to guidelines strictly: *“hundreds of South Asians across Croydon. Everybody is following strictly*.*”* (P4) or *“As far as I know, um, my community, uh, we are from the part, where in India, I see everybody is just taking things seriously and behaving responsibly”* (P13). This adherence, this group efficacy—like self-efficacy—was enabled by their position as an ‘integrated immigrant’: they drew support from their community, religion, and family living arrangements while also espousing the rule of law and respect for UK institutions.

## Discussion

We find that the environment of health and identity threat characterised by awareness of BAME COVID risk was managed by the participant group by asserting efficacy that was congruent with identity construction. The integrated immigrant identity asserted by our participants was inextricable from self-, group- and institutional efficacies. These findings suggest that the psychological processes of the deployment of efficacy resources and of identity management are inextricably linked, extending both the health and identity literatures related to threat management. These findings contribute theoretically to the investigation of efficacy in health behaviours, and point to lessons to be taken from the construction of an identity that successfully retains important cultural elements, while incorporating efficacies essential for the promotion of health behaviours. This constructed identity, however, asserted the heterogeneity of the BAME group. Other members of the BAME community were positioned as less efficacious due to their less integrated identities, highlighting intragroup stigmatisation as an unintended consequence of treating the BAME community as a homogeneous one.

Our findings are in line with identity threat literature that predicts that those who are involuntarily categorised will distance themselves from the ascribed label and align themselves with a non-threatened group, will assert the heterogeneity of the group, and assert their personal attributes (Branscombe et al., 1999; Doosje et al., 1999; Ellemers et al., 2002). Participants resisted negative social categorisation by challenging the implication that they were representative of the ethnic or South Asian community. Instead, they opted to construct an alternative identity to the one presented to them. The participants, through their discussions of threat response, constructed a sub-group of the South Asian community, whose members identified with both the ethnic and White majority groups. They constructed an identity that incorporated specific cultural norms from their own culture as well as those that align more closely with their adopted country, creating an integrated immigrant identity specific to this group. Participants in this study therefore expressed their efficacy within a context of tension between their South Asian culture and norms of the majority British population expressed through an ‘integrated immigrant’ identity.

Through this ‘integrated immigrant’ identity, we found that—in line with Cho and Kuang (2015), self-, group-, and institutional efficacies were all drawn upon to manage threat response. Participants asserted efficacies that both aligned with the white British majority suggesting that cultural assimilation could lead to lifestyle changes, self-reliance (an assertion of self-efficacy) and faith in the NHS and UK government (an assertion of institutional efficacy), as well as group efficacy according to their constructed identity. The participant group directly asserted their beliefs in self-responsibility, a belief that is integral to the cornerstone UK belief in social mobility (endorsed by 85% of UK ISSP respondents in 2010). Such social mobility beliefs are typical of highly skilled immigrants, similar to our participants (Lumpe, 2019), suggesting the importance of immigrant economic status in response to this health threat, not just in terms of economic resources but also in self-efficacy beliefs. This finding is in contrast to work that indicates that South Asian and other ethnic minorities are less likely to espouse self-efficacy in the context of health threat (Hendy et al., 2019; Vandrevala et al., under review). The assertion of institutional efficacy appeared in the widely held views of equal access to healthcare and the endorsement of the UK government’s virus response. Like Cho et al. (2020), we too find that institutional efficacy is not inversely related to derogation of the outgroup as our group of participants represented their intra-ethnic outgroups as responsible for COVID-19 violations. It is possible that support for institutional efficacy may speak to support for the status quo by those who are beneficiaries of the current system. In our data, it was a device that allowed a minority group under threat to align itself with the majority group, thus playing a role in identity construction.

### Implications for practice

Group efficacy, a key element of collective mobilisation (Van Zomeren et al., 2008) reflected participants’ integrated immigrant South Asian identity. The group was presented not only in terms of the British norms expressed through self- and institutional efficacy, but also through the South Asian cultural norms of family support (including multi-generational living), community, and religion. As such, it highlights values and norms that are important to these participants in their negotiations between their ethnic and national identities. The centrality of these features is emphasised in the current context as certain of these cultural facets could also be framed as risk-enhancing during the pandemic. Multi-generational living, for example, simply through its nature of having more people sharing living quarters means that the risk of exposure to the virus is higher. Regardless, this high-status group was able to accommodate this cultural aspect as a core part of their identity, extolling the benefits of the arrangements, while acknowledging the risk. Rather than framing culturally specific traditions and customs (such as, multi-generational living) as risky, our study suggests that there are lessons to be gained regarding the successful integration of cultural identity and efficacies as opposed to the stigmatising narrative being used to position ethnic community customs and living arrangement as posing additional risk. Health messages should explicitly consider cultural norms, ensure they promote services that are accessible and do not disadvantage ethnic community and consider beliefs, attitudes and behaviour, socio-cultural factors and the range of influences and drivers of behaviour which may, at times, differ not only from White British communities, but also within their ethnic group.

In addition to structuring efficacy, the construction of this identity also provides insight into the commonalities within the South Asian community that the participant group values, and those features that they believe differentiates them from their ethnic group— observations that have implications for group mobilisation related to health behaviours. Commonalities were noted in the area of religious observation and the more social structure of the community. The identification of commonalities is particularly important in a situation of societal risk such as the COVID-19 pandemic. Group identification and perceived group norms are key for messaging to lead to mobilisation and enactment of pro-health behaviours. More effective group-directed messaging reflects an individual’s understanding of themselves as a member of the group (Drury et al., 2020; Reicher and Hopkins, 2001) and their role in recognition of threat and adherence to the required public health behaviours.

More generally, however, the assertions of self-, group-, and institutional efficacy reflected perceived differences between the participant group and the broader South Asian group; differences primarily related to the in-group’s specific areas of integration: diet, exercise and personal hygiene practices by the participant group was distinguished from less integrated South Asians. The importance of religion to social life was recognised, but differing priorities regarding religious rules and the British ‘rule of law’ distinguished between the participant group and their South Asian out-group. Such observations of self-defined groups are important for understanding how these participants define their social selves, and therefore the social identities that are relevant for connecting communities, for public service messaging, and for group mobilisation. These findings speak not only to the limitations of addressing South Asians as a community, but also to the intragroup stigmatization that can accompany categorisation threat. (Abrams et al., 2021) highlight that some groups (often those that are more disadvantaged) can become targets of blame or stigmatization associated with their perceived risky behaviour or their vulnerability to the virus, or both. Emphasis shifts from depicting particular groups sympathetically because of their high personal vulnerability to infection to scrutinising those groups’ behaviours in the quest to assign blame for not respecting the social distancing guidelines and these can result in emerging tensions between newly salient social groups. Furthermore, they suggest the need for messaging and guidance tailored to ethnic minority communities with varying levels of association and communication with ‘majority British’ culture and institutions to avoid unintended stigmatisation.

### Limitations

These findings are based on a particular subset of the BAME community—middle class South Asians who were primarily Indian and Hindu. This study was based on a participant group that reflected only one particular segment of the BAME community who occupies a higher economic status and level of integration than other members of the BAME community. This group was able to exercise their efficacies through their identities as an integrated immigrant. BAME community members for whom this group mobility is less available may be more likely to ‘turn-in’ towards their own group in the same threat context, and may engage in increased self-stereotyping and outgroup derogation (Branscombe et al., 1999; Ellemers et al., 2002). Lower economic status and less integrated groups are therefore unlikely to have the same identity/efficacy expression available to them, which could plausibly bring about greater resistance to public health messaging. Future work may employ our theoretical intertwining of efficacy and identity to explore such relationships in other sub-populations. Comparative work would be particularly useful in this area. Rather than framing ethnicity as a key variable in COVID-19 risk in public and policy deliberations, it would be more appropriate to consider pre-existing social, environmental and economic inequality that have been exhibited during the pandemic (Marmot et al., 2020) in health message framing and recovery from the pandemic.

## Conclusion

Our findings have implications for theory and practice regarding health behaviours in the BAME community. Specifically, it provides evidence of negative implications of treating the BAME community as a homogenous group. The work also sheds light on the influence of class, group efficacy, and institutional trust in promoting attitudes towards protective health behaviours. These insights are particularly valuable in addressing public health threats such as vaccine hesitancy in the wider BAME group, but may also be applied to individual health behaviours, such as engaging with COVID-19 health protecting behaviours and test-trace and isolate systems.

## Data Availability

The raw data can be made available upon request

---

**Table 1** *Participant information*

readers may contact corresponding author for access to data

